# Machine learning vs. traditional regression analysis for fluid overload prediction in the ICU

**DOI:** 10.1101/2023.06.16.23291493

**Authors:** Andrea Sikora, Tianyi Zhang, David J. Murphy, Susan E. Smith, Brian Murray, Rishikesan Kamaleswaran, Xianyan Chen, Mitchell S. Buckley, Sandra Rowe, John W. Devlin, the MRC-ICU Investigator Team

## Abstract

**Background:** Fluid overload, while common in the ICU and associated with serious sequelae, is hard to predict and may be influenced by ICU medication use. Machine learning (ML) approaches may offer advantages over traditional regression techniques to predict it. We compared the ability of traditional regression techniques and different ML-based modeling approaches to identify clinically meaningful fluid overload predictors.

**Methods:** This was a retrospective, observational cohort study of adult patients admitted to an ICU ≥ 72 hours between 10/1/2015 and 10/31/2020 with available fluid balance data. Models to predict fluid overload (a positive fluid balance ≥10% of the admission body weight) in the 48-72 hours after ICU admission were created. Potential patient and medication fluid overload predictor variables (n=28) were collected at either baseline or 24 hours after ICU admission. The optimal traditional logistic regression model was created using backward selection. Supervised, classification-based ML models were trained and optimized, including a meta-modeling approach. Area under the receiver operating characteristic (AUROC), positive predictive value (PPV), and negative predictive value (NPV) were compared between the traditional and ML fluid prediction models.

**Results:** A total of 49 of the 391 (12.5%) patients developed fluid overload. Among the ML models, the XGBoost model had the highest performance (AUROC 0.78, PPV 0.27, NPV 0.94) for fluid overload prediction. The XGBoost model performed similarly to the final traditional logistic regression model (AUROC 0.70; PPV 0.20, NPV 0.94). Feature importance analysis revealed severity of illness scores and medication-related data were the most important predictors of fluid overload.

**Conclusion:** In the context of our study, ML and traditional models appear to perform similarly to predict fluid overload in the ICU. Baseline severity of illness and ICU medication regimen complexity are important predictors of fluid overload.

## INTRODUCTION

Fluid overload, a frequent and unintended consequence of the resuscitation process in critically ill adults may result in increased rates of acute kidney injury and invasive mechanical ventilation initiation, prolonged intensive care unit (ICU) stay, and mortality (1, 2). Timely de-resuscitation to remove excess fluid is associated with improved outcomes (3-6). While the predictors of volume responsiveness are well-established (7, 8), the predictors for ICU fluid overload remain unclear (7, 8). Development of rigorous fluid overload prediction algorithms could shorten the time to the implementation of fluid overload mitigation strategies [e.g., concentration of intravenous (IV) fluid products, discontinuation of maintenance fluids, administration of diuretics] and improve outcomes.

Non-diuretic ICU medication use may affect fluid overload risk; preliminary data suggests the medication regimen complexity-ICU (MRC-ICU) score is associated with both fluid overload and fluid balance (9). This score has also been shown to predict mortality and length of stay and also the medication interventions needed to optimize a patient’s pharmacotherapy regimen (10-17). Therefore, quantifying patient-specific, medication-related data is likely an important consideration in the prediction of fluid overload in critically adults (2, 18, 19).

Event prediction in the ICU remains a perennial area of research given the many challenges that exist for clinicians to accurately predict clinical outcomes in the highly complex and dynamic critical care environment (20, 21). Artificial intelligence and machine learning techniques have been proposed as a method to improve ICU clinical outcome prediction given their unique ability to handle multi-dimensional problems and identify novel patterns within the vast troves of continuously-generated patient data (19, 22-24). However, to some ICU clinicians, the use of artificial intelligence/machine learning approaches to predict clinical events may have a ‘black-box effect,’ which can ultimately preclude implementation. The rigorous evaluation of whether artificial intelligence-based approaches predict clinical events better than traditional regression models (or clinical expertise alone) remains a key question in critical care practice (25-29).

In this study, we sought to compare the ability of machine learning approaches to traditional regression models to predict fluid overload and the individual predictors for its occurrence in critically ill adults. We hypothesized that advanced machine learning techniques perform better than traditional regression models to predict fluid overload and that the predictors for fluid overload identified through machine learning approaches may be different.

## METHODS

We conducted a retrospective, observational study of adults admitted ICUs at the University of North Carolina Health System (UNCHS), an integrated health system, who had fluid overload data available. The protocol for this study was approved with waivers of informed consent and HIPAA authorization granted by UNHCS Institutional Review Board (approval number: (Project00001541); approval date: October 2021). Procedures followed in the study were in accordance with the ethical standards of the of the UNHCS Institutional Review Board and the Helsinki Declaration of 1975, as most recently amended (30). The reporting of this study adheres to the STrengthening and reporting of OBservational data in Epidemiology statement (31).

### Population

A random sample of 1,000 adults (≥18 years) admitted to an ICU at UNCHS between October 2015 and October 2020 was generated. Patients on their index ICU admission with fluid balance data available for the first 72 hours were included (**Supplemental Digital Content (SDC) Figure 1**). Patients were excluded if the admission was not their index ICU admission.

**Figure 1.** Feature importance for presence of fluid overload prediction with XGBoost

### Data Collection and Outcomes

De-identified UNCHS electronic health record (EHR) data (Epic Systems, Verona, WI) housed in the Carolina Data Warehouse (CDW) was extracted by a trained CDW data analyst. The primary outcome was the presence of fluid overload at the 48-72 hours (i.e., day 3) after ICU admission. Fluid overload was defined as a positive fluid balance in milliliters (mL) greater than or equal to 10% of the patient’s admission body weight in kilograms (kg) (2, 32). For example, a patient with a body weight of 100kg at ICU admission having a positive fluid balance at 72 hours of 12,000 mL (or 12kg) would be considered to have fluid overload. A secondary outcome was the amount of fluid overload as a function of body weight. For example, the aforementioned patient would have a fluid overload amount of 12%.

Following a literature review, and through investigator consensus, potential predictor variables for fluid overload were defined (2, 33-36). A total of potential 28 predictors were identified: 1) *ICU baseline*: age ≥ 65 years, sex, admission to a medical (vs. surgical) ICU, primary ICU admission diagnosis (i.e., cardiac, chronic kidney disease, heart failure, hepatic, pulmonary, sepsis, trauma), and select co-morbidities (i.e., chronic kidney disease, heart failure); 2) *24 hours after ICU admission*: APACHE II and SOFA score (using worst values in the 24 hour period), use of supportive care devices (i.e., renal replacement therapy, invasive mechanical ventilation), serum laboratory values (i.e., albumin < 3 mg/dL, bicarbonate < 22 mEq/L or > 29 mEq/L, chloride ≥ 110 mEq/L, creatinine ≥ 1.5 mg/dL, lactate ≥ 2 mmol/L, potassium ≥ 5.5 mEq/L, sodium ≥ 148 mEq/L or < 134 mEq/L), fluid balance (mL), and presence of acute kidney injury (as defined by need for renal replacement therapy or serum creatinine greater than or equal two times baseline); 3) *Medication data at 24 hours*: MRC-ICU score, vasopressor use in the first 24 hours, use of continuous medication infusions, and the number of continuous medication infusions.

### Data Analysis

#### Data Missingness

Due to the hypothesis-generating nature of our study and the lack of published data on ICU fluid overload prediction, no attempt was made to estimate a study sample size. Multiple imputation (10) imputations per variable was applied for all missing data (see **Supplemental Digital Content (SDC)**).

#### Machine Learning Models

We employed Random Forest, SVM and XGBoost for the task of modeling the presence of fluid overload (37-39). During the model training on each of the ten imputed training sets, 5-fold cross validation was applied for Random Forest, SVM and XGBoost to choose the hyperparameters for these machine learning models. With the optimal hyperparameters, the models were fitted again on the corresponding imputed training set. Predictions for probability of fluid overload were made on each of the ten imputed testing sets using the corresponding optimal model. For Random Forest, two hyperparameters were tuned (number of trees and number of variables randomly sampled as candidates at each split). For SVM, linear kernel and cost of constraints violation were tuned. For XGBoost, two hyperparameters were tuned (maximum depth of a tree and maximum number of boosting iterations). For each model, there were ten different imputed test sets that then generated ten different predictions. These predictions of the probability for fluid overload were averaged as the final prediction.

For the degree of fluid overload, we built models with the amount of fluid overload at 72 hours. Since this is a continuous variable, we employed their regression of the above machine learning models: Random Forest regression, SVM regression, and XGBoost regression. For XGBoost, feature importance was measured as the frequency a feature was used in the trees. For Random Forest, feature importance was measured by mean decrease in node impurity. Because ten different models were used on each imputed dataset, ten different feature importance lists were generated for each. A subsequent analysis modeling fluid overload as a continuous variable (percent of net milliliters of fluid by body weight) instead of dichotomous presence or absence of fluid overload) was performed (see **SDC**).

#### Traditional Regression Models

991After multiple imputation, each of the ten completed datasets was split into training data and testing data using an 80:20 ratio. Subsequently, a full logistic regression model was built for the presence of fluid overload for each of the ten complete training sets. We then applied backward elimination to select the final model. The initial set of variables for the variable selection were determined by the significance of variables in the ten full models. We built our linear regression models so that the degree of fluid overload was similar to that of the ten completed training sets. In order to compare these models with the MRC-ICU only model, we also built logistic regression and linear regression models with MRC-ICU as the sole predictor in the ten training sets. After model fitting, model fits were pooled using Rubin’s method (40). Using the pooled models, odds ratios (OR) and their 95% confidence intervals (CI) were reported.

## RESULTS

A total of 49 (12.5%) of the 391 included patients had fluid overload on ICU day 3. The degree of day 3 fluid overload was significantly greater in the fluid overload (vs non overload) patients (16.6% vs 2.2%, p < 0.01). Overall, the mean APACHE II score was 15.7 ± 6.6, mean SOFA score was 8.3 ± 3.3, and MRC-ICU score was 11.8 ± 8.7. A significantly greater proportion of fluid overload patients (vs. those without) had an elevated serum lactate ≥ 2 mmol/L (32.7% vs. 14.9%, p = 0.01) and AKI (28.6% vs. 10.5%, p < 0.001) at 24 hours and positive fluid balance (1,840 mL vs. 390 mL, p < 0.001) on ICU day 3. All model covariates are summarized in **Table 1**. At ICU day 3, patients with fluid overload (vs those without) were more likely to be dead (20.4% vs. 7.3%, p = 0.01), have AKI (34.7% vs. 15.8%, p < 0.001), and remain on mechanical ventilation (12.7% vs. 4.2%, p = 0.05).

**Table 1.** Study cohort characteristics by presence of fluid overload within 72 hours of ICU admission

|  | All<br>(n = 391) | Fluid Overload<br>(n = 49) | No Fluid Overload<br>(n = 342) | p-value |
| --- | --- | --- | --- | --- |
| **ICU Baseline** |  |  |  |  |
| Age ≥ 65 years | 202 (51.7) | 19 (38.8) | 183 (53.5) | 0.08 |
| Male sex | 213 (54.5) | 23 (46.9) | 190 (55.6) | 0.33 |
| Chronic comorbidities |  |  |  |  |
| Chronic kidney disease | 13 (3.3) | 1 (2.0) | 12 (3.5) | 0.06 |
| Heart failure | 19 (4.9) | 2 (4.1) | 17 (4.9) | 0.06 |
| Admission to medical ICU | 156 (39.9) | 24 (48.9) | 132 (38.6) | 0.22 |
| Primary ICU Admission Diagnosis |  |  |  |  |
| Cardiac | 81 (20.7) | 3 (6.1) | 78 (22.8) | 0.06 |
| Chronic kidney disease | 13 (3.3) | 1 (2.0) | 12 (3.5) |  |
| Hepatic | 6 (1.5) | 1 (2.0) | 5 (1.5) |  |
| Pulmonary | 58 (14.8) | 8 (16.3) | 50 (14.6) |  |
| Sepsis/septic shock | 29 (7.4) | 7 (14.3) | 22 (6.4) |  |
| Trauma | 10 (2.6) | 3 (6.1) | 7 (2.0) |  |
| **24 hours after ICU admission** |  |  |  |  |
| Severity of illness, mean (SD) |  |  |  |  |
| APACHE II Score | 15.7 (6.6) | 17.5 (7.0) | 15.4 (6.6) | 0.06 |
| SOFA Score | 8.3 (3.3) | 9.9 (4.6) | 8.2 (3.1) | 0.07 |
| Supportive devices |  |  |  |  |
| Any renal replacement therapy | 5 (1.3) | 1 (2.0) | 4 (1.2) | 1.00 |
| Any mechanical ventilation | 140 (35.8) | 21 (42.9) | 119 (34.8) | 0.53 |
| Serum laboratory values |  |  |  |  |
| Albumin $<$3 mg/dL | 88 (22.5) | 18 (36.7) | 70 (20.5) | 0.02 |
| Bicarbonate $<$ 22 mEq/L | 74 (18.9) | 14 (28.6) | 60 (17.5) | 0.16 |
| Bicarbonate $>$ 29 mEq/L | 64 (16.4) | 6 (12.2) | 58 (16.9) | |
| Creatinine ≥ 1.5 mg/dL | 28 (7.2) | 7 (14.3) | 21 (6.1) | 0.02 |
| Chloride ≥ 110 mEq/L | 125 (31.9) | 19 (38.8) | 106 (30.9) | 0.33 |
| Potassium ≥ 5.5 mEq/L | 19 (4.9) | 5 (10.2) | 14 (4.1) | 0.12 |
| Lactate ≥ 2 mmol/L | 67 (17.1) | 16 (32.7) | 51 (14.9) | 0.01 |
| Sodium ≥ 148 mEq/L | 22 (5.6) | 6 (12.2) | 16 (4.7) | 0.01 |
| Sodium $<$134 mEq/L | 33 (8.4) | 4 (8.1) | 29 (8.5) | |
| Fluid balance (mL), mean (SD) | 570 (1960) | 1840 (301) | 390 (168) | $<$0.001 |
| Acute kidney injury | 50 (12.8) | 14 (28.6) | 26 (10.5) | $<$ 0.001 |
| Medications |  |  |  |  |
| MRC-ICU, mean (SD) | 11.8 (8.7) | 13.4 (8.4) | 11.5 (8.7) | 0.06 |
| Any vasopressor | 119 (30.4) | 16 (32.6) | 103 (30.1) | 0.85 |
| Any continuous infusions | 249 (63.6) | 34 (69.3) | 215 (62.8) | 0.47 |
| Infusions / patient, mean (SD) | 2.29 (3.3) | 1.98 (2.2) | 2.33 (3.4) | 0.35 |

Data are presented as n (%) unless otherwise stated

Among the machine learning models, XGBoost demonstrated the highest AUROC (0.78) compared to SVM (0.69) and RF (0.76) and was associated with a PPV of 0.27 and NPV of 0.94. Notably, all models tested at relatively poor PPV. In comparison, stepwise logistic regression had an AUROC of 0.70, PPV 0.26, and NPV 0.94. Full results are reported in **Table 2**, and AUROC curves for all models are provided in **SDC Supplemental Figure 2**. Results of the full logistic regression are reported in **SDC Supplemental Table 1**. Stepwise regression resulted in a more parsimonious model (7 variables vs. 31 variables) but demonstrated similar performance to the machine learning models (**SDC Supplementary Table 2**). In the stepwise regression, presence of sepsis, male sex, the SOFA score at 24 hours, and the 24 hour serum sodium and bicarbonate comprised the stepwise regression model (**Table 2**). In an analysis of MRC-ICU as a single predictor for fluid overload, the model had an AUROC of 0.74 (0.60-0.84), sensitivity 0.62 (0.35-0.85), specificity 0.70 (0.63-0.77), PPV 0.16 (0.08-0.27), and NPV 0.96 (0.90-0.98).

**Table 2.** Performance of presence of fluid overload prediction models, mean (confidence interval)

|  | **AUROC** | **Sensitivity** | **Specificity** | **PPV** | **NPV** |
| --- | --- | --- | --- | --- | --- |
| *Traditional regression* |  |  |  |  |  |
| **All variables** | 0.70 (0.53, 0.82) | 0.43 (0.19, 0.70) | 0.85 (0.79, 0.89) | 0.20 (0.08, 0.37) | 0.94 (0.89, 0.97) |
| **Stepwise Selected Regression** | 0.70 (0.52, 0.82) | 0.43 (0.19, 0.70) | 0.89 (0.84, 0.93) | 0.26 (0.11, 0.47) | 0.94 (0.90, 0.97) |
| *Supervised machine learning models* |  |  |  |  |  |
| **Random Forest** | 0.76 (0.62, 0.86) | 0.56 (0.29, 0.80) | 0.8571 (0.80, 0.90) | 0.25 (0.12, 0.43) | 0.95 (0.91, 0.98) |
| **Support Vector Machine** | 0.69 (0.51, 0.82) | 0.50 (0.24, 0.75) | 0.82 (0.76, 0.88) | 0.21 (0.09, 0.36) | 0.94 (0.90, 0.97) |
| **XGBoost** | 0.78 (0.62, 0.87) | 0.37 (0.15, 0.64) | 0.91 (0.86, 0.94) | 0.27 (0.10, 0.50) | 0.94 (0.89, 0.97) |

AUROC: area under the receiver operating characteristic; PPV: positive predictive value; NPV: negative predictive value

Feature importance graphs were plotted for XGBoost (**Figure 1**), RF (**SDC Supplemental Figure 3)** and SVM (**SDC 5 Supplemental Figure 4)**. Among the 10 different feature importance lists generated for each model, differences between top features were noted. For example, for two of the machine learning models, XGBoost (**Figure 2**) and RF, the top five most important features were fluid balance at 24 hours, SOFA score at 24 hours, MRC-ICU at 24 hours, APACHE II at 24 hours, and the number of continuous infusions at 24 hours. While the stepwise regression model found fluid balance at 24 hours and APACHE II at 24 hours to be top features, the SOFA score at 24 hours, the MRC-ICU at 24 hours and the number of continuous infusions were not found to be model features.

**Figure 2.** Most common features for presence of fluid overload prediction with XGBoost imputations

The full regression results for predicting the amount of fluid overload at 72 hours are reported in **SDC Supplemental Table 3**. For stepwise regression, twelve variables were included with fluid balance, laboratory values, and severity of illness being significant predictors (**SDC Supplemental Table 4**). All models demonstrated similar performance as measured by MSE (**SDC Supplemental Table 5**). Feature importance graphs are presented **in SDC Supplemental Figures 5-7**).

## DISCUSSION

Although machine learning models have been shown to outperform traditional regression models in a variety of settings (41, 42), the potential benefits of machine learning in critical care remain an open field of exploration, in part due to a current lack of rigorous comparison in high quality ICU datasets (27, 43, 44). Our analysis represents the first published comparison of machine learning approaches with traditional regression methods to predict fluid overload using a novel dataset with granular medication data.

We report that machine learning and logistic regression analyses demonstrate a similar predictive power to identify patients with fluid overload on day 3 of their ICU stay. Although use of machine learning did not appear to improve predictive performance over regression analysis, it expanded the number of variables critical to fluid overload prediction and highlights the importance of further artificial intelligence-based exploration in this area. This analysis of individual predictors may help bedside clinicians better understand how the machine learning models work and may help overcome their ‘black box’ hesitancy to trust machine learning-generated results (45, 46). For example, feature importance graphs for the machine learning analyses found complexity of the daily ICU medication regimen (i.e., MRC-ICU score), which includes the number of intravenous medication infusions (the primary method to administer medications in this population and a primary source of fluids to a patient), to be an important contributor to fluid overload. In comparison, in the traditional multivariable regression, the MRC-ICU score was not associated with fluid overload. This may be because machine learning analyses better account for severity of illness and the response of clinicians to respond to this severity by administering more medication infusions leading to a more complex daily medication regimen; however, the methods applied, including feature importance, preclude causal inference at this juncture. As such, our results highlight the unique power of machine learning to identify complex relationships that can be further elucidated via machine-learning based causal inference modeling and other designs aimed at causation (2, 18).

Optimizing fluid management (or fluid stewardship) has been previously defined by the ROSE model of Resuscitation, Optimization, Stabilization, and dE-resuscitation (33). After an initial 24-48 hour period characterized by overt volume resuscitation (e.g., a crystalloid bolus) and IV medication initiation (e.g., antibiotics), and the associated fluid administration, the care priority shifts from volume administration to volume removal. While comprehensive fluid stewardship management strategies including reduced fluid use and diuretic administration can effectively reduce fluid overload and its sequelae, they are often deployed too late (1, 2). Interstingly, some reports have indicated ‘hidden fluids’ (defined as blood products, enteralnutrition, flushes, and intravenous medications) were significantly associated with the development of fluid overload. While in critical illness many of these ‘hidden fluids’ are necessary (e.g., blood products), given that intravenous medications account for over 40% of total fluid intake in this analysis, interventions such as concentrating intravenous medications, employing oral formulations when feasible, careful evaluation of maintenace fluids, and antibiotic de-escalation are potoentially still viable even in high illness severity that can reduce this complication. However, weighing risks and benefits associated with these interventions in context may yet be aided by more quantitative prediction data (50, 51). Overall, de-resuscitation and fluid stewardship can be deceptively complex (47). In a patient with shock, balancing the dueling forces of volume responsiveness assessment and timely volume resuscitation with the risks associated with fluid overload represents a highly complex Goldilocks scenario that requires clinicians to have high clinical precision, essentially pivoting ‘on a dime’, from a strategy of aggressive volume expansion to one of rapid volume removal (34, 48, 49).

Despite the complexities of this decision process, limited prediction tools for fluid overload are available to assist clinicians at the ICU bedside. As such, real-time recognition identifying when to make the shift from resuscitation to de-resuscitation has the potential to improve bedside management. However, to go beyond the hourly assessment of ‘Ins and Outs’ would require accurate prediction of future fluid overload risk and the adverse events associated with it, in the time-dependent context of intervention delivery (e.g., diuretics). In such a scenario, an algorithm would be able to accurately interpret a septic patient who is 3 liters positive 24 hours after fluid resuscitation initiation as being in a ‘green zone’ (i.e., appropriately resuscitated). However, 24 hours later, if the same patient is 4 liters positive while off vasopressors and with down-trending sepsis markers the algorithm could alert clinicians that the patient is now in a ‘yellow zone’ where interventions like diuretic therapy and fluid reductions are required to reduce acute kidney injury and intubation risk. This type of real-time predictive capability could support continuous clinician decision-making but requires evaluation outside the scope of our current study.

Fluid overload also presents an important test case for exploring and adapting artificial intelligence methods to ICU problems, particularly those related to ICU medication use. Fluid overload represents a uniquely *intervenable event* in the ICU. Intervenable events share three key characteristics: they are *predictable, preventable*, and otherwise associated with *poor* outcomes. The results of our study, and others, indicate that fluid overload can be *predicted* with modeling of some kind, especially given its ability to be quantitatively defined (50-52). Fluid overload has been associated with poor outcomes including acute kidney injury, delirium, poor respiratory outcomes, prolonged length of stay, and potentially increasing mortality (2, 35, 53-56). Evidence demonstrates the timely recognition and management of fluid overload is feasible and is associated with reduced mortality and time in the ICU (3, 57-60). Notably, fluid stewardship has been adapted by critical care pharmacists as key component of comprehensive medication management (5, 6, 60). As such, these results may support other investigations as they identify patients in whom it is safe to initiate de-resuscitation or importantly never needed that degree of fluid volume initially and at the bedside may prompt clinicians to be more targeted in therapies initiated or aggressive in curtailing early ‘hidden’ fluids to avoid the complications of fluid overload and/or the need for a highly interventional period of de-resuscitation (e.g., diuretics, dialysis). Artificial intelligence may be particularly well suited to bolster these efforts, and thus while feature importance analyses cannot provide foundation for causal inference, they may guide such future investigations.

Our study has limitations. Our patient sample may have been too small to demonstrate superiority of the machine learning approaches compared to traditional regression, and no validation in a separate, external dataset was undertaken at this juncture (61). Bias may exist due to which patients had fluid balance data available. Other predictors for fluid overload not included in our models may exist (62). By relying on prediction data derived in the first 24 hours of ICU admission, we did not fully capture the dynamic nature of critical illness over the entire three day ICU period before fluid overload occurred. Future time-dependent evaluations of changing features employing unsupervised learning techniques may yield novel insights.

## CONCLUSION

Fluid overload is an important, intervenable event in the ICU population. Incorporation of medication-related variables and artificial intelligence has demonstrated promise to improve prediction that may ultimately guide timely intervention and mitigation of this ICU complication; however, comparative advantages over traditional modeling techniques may remain warranted.

## Supporting information

Supplemental Content

## Data Availability

All data produced in the present study are available upon reasonable request to the authors

## Acknowledgements

Data acquisition were supported by NC TraCS, funded by Grant Number UL1TR002489 from the National Center for Advancing Translations Sciences at the National Institutes of Health, and Data Analytics at the University of North Carolina Medical Center Department of Pharmacy.

## Declarations

### Ethical Approval

The protocol for this study was approved with waivers of informed consent and HIPAA authorization granted by UNHCS Institutional Review Board (approval number: (Project00001541); approval date: October 2021).

### Author Contributions

A.S. was responsible for project execution, design, and initial manuscript writing. J.D., D.M., and R.K. provided critical revisions of manuscript, data interpretation, and senior level oversight. M.Y., T.Z, and X.C. handled data pre-processing and analysis (M.Y., T.Z.) and methodology support and data interpretation (X.C., R.K.). B.M. served as site coordinator for all data validation and procurement as well as manuscript revisions and data interpretation. S.S., M.B., and S.R. provided clinical interpretation, results interpretation, and manuscript revision.

### Availability of data & materials

The datasets used and/or analyzed during the current study available from the corresponding author on reasonable request.

