## Supplemental Content for "Machine learning vs. traditional regression analysis for fluid overload prediction in the ICU"

**Supplementary Material**

**Methods**

**Supplemental Table 1.** Full regression model for presence of fluid overload at 72 hours

**Supplemental Table 2.** Stepwise regression for final model to predict presence of fluid overload

**Supplemental Table 3.** Full regression model for amount of fluid overload at 72 hours

**Supplemental Table 4.** Stepwise regression for final model to predict fluid overload amount

**Supplemental Table 5.** Performance of fluid overload prediction models for amount of fluid overload

**Supplemental Figure 1.** Consort diagram

**Supplemental Figure 2.** AUROC Curve for Fluid Overload at 72 hours

**Supplemental Figure 3.** Most common features for presence of fluid overload prediction with Random Forest imputations

**Supplemental Figure 4.** Most common features for presence of fluid overload prediction with Support Vector Machine imputations

**Supplemental Figure 5.** Most common features for amount of fluid overload prediction with XGBoost imputations

**Supplemental Figure 6.** Most common features for amount of fluid overload prediction with Random Forest imputations

**Supplemental Figure 7.** Most common features for amount of fluid overload prediction with Support Vector Machine imputations

**Methods**

*Data Missingness:* Variables were excluded if the corresponding missing proportions were over 30% prior to imputation, with the exception of SOFA score. For continuous variables [including APACHE II, SOFA, fluid balance (mL), and amount of fluid overload], linear regression was applied for imputation. During the imputation process, logistic regression was used for binary variables and polytomous logistic regression was used for multi-level variables. Descriptive statistics on the data before multiple imputation were calculated, and clinical characteristics between those patients with and without fluid overload were compared using either Student *t* test or Chi-square test, as appropriate. A p-value of less than 0.05 was considered statistically significant.

*Amount of fluid overload analysis:* All methodology for calculating percent fluid overload as a continuous variable was similar except for appropriate changes to manage a continuous variable including the use of linear regression (instead of logistic) and the use of Random Forest regression, SVM regression, and XGBoost regression instead of Random Forest classifier, SVM classifier, and XGBoost classifier. Finally, performances were measured using mean squared error (MSE) calculated on each imputed testing set and then averaged as the final MSE. To compare performance of the models, a series of values were calculated including for the models with classification task: area under the receiver operating characteristic (AUROC), positive predictive value (PPV), negative predictive value (NPV), specificity, and sensitivity. For regression models, mean square error (MSE) was used to measure the model performance.

**Supplemental Table 1.** Full regression model for presence of fluid overload at 72 hours

|  | **Univariate** | | | **Multivariate** | | |
| --- | --- | --- | --- | --- | --- | --- |
| **Variable** | **Odds Ratio** | **95% CI** | **p-value** | **Odds Ratio** | **95% CI** | **p-value** |
| **ICU Baseline** | | | | | | |
| Age ≥ 65 years old; mean (SD) | 0.73 | 0.45, 1.18 | 0.19 | 0.57 | 0.29, 1.10 | 0.09 |
| Sex (male) | 0.51 | 0.31, 0.83 | 0.00 | 0.48 | 0.26, 0.88 | 0.01 |
| *Relevant chronic conditions* | | | | | | |
| Chronic kidney disease | 0.42 | 0.05, 3.07 | 0.39 | 0.26 | 0.02, 3.07 | 0.28 |
| Heart failure | 0.80 | 0.18, 3.55 | 0.77 | 3.05 | 0.53, 17.55 | 0.20 |
| Admission to medical ICU | 1.17 | 0.69, 1.97 | 0.53 | 0.92 | 0.41, 2.07 | 0.84 |
| *Primary ICU Admission Diagnosis* | | | | | | |
| Cardiac | 0.42 | 0.16, 1.07 | 0.07 | 0.65 | 0.18, 2.28 | 0.49 |
| Chronic kidney disease | 0.42 | 0.05, 3.07 | 0.39 | 0.26 | 0.02, 3.07 | 0.28 |
| Hepatic | 1.05 | 0.14, 7.86 | 0.95 | 0.43 | 0.02, 6.59 | 0.54 |
| Pulmonary | 1.12 | 0.56, 2.24 | 0.73 | 1.20 | 0.47, 3.06 | 0.68 |
| Sepsis/septic shock | 4.22 | 2.17, 8.21 | 0.00 | 3.50 | 1.51, 8.11 | 0.00 |
| Trauma | 1.19 | 0.41, 3.40 | 0.75 | 0.46 | 0.11, 1.98 | 0.30 |
| **24 hours after ICU admission** | | | | | | |
| *Severity of illness, using worst values recorded* | | | | | | |
| APACHE II Score | 1.11 | 1.07, 1.15 | 0.00 | 1.07 | 0.99, 1.16 | 0.06 |
| SOFA Score | 1.31 | 1.21, 1.42 | 0.00 | 1.20 | 0.99, 1.44 | 0.05 |
| *Supportive care devices* | | | | | | |
| Renal replacement therapy | 0.85 | 0.10, 6.79 | 0.88 | 0.17 | 0.01, 2.00 | 0.16 |
| Invasive mechanical ventilation | 2.95 | 1.89, 4.61 | 0.00 | 0.67 | 0.25, 1.77 | 0.42 |
| *Laboratory values (serum) and flowsheet values* | | | | | | |
| Bicarbonate < 22 mEq/L | 0.37 | 0.22, 0.62 | 0.00 | 0.59 | 0.31, 1.13 | 0.11 |
| Bicarbonate > 29 mEq/L | 0.22 | 0.09, 0.53 | 0.00 | 0.36 | 0.12, 1.10 | 0.07 |
| Creatinine ≥ 1.5 mg/dL | 2.48 | 1.25, 4.91 | 0.00 | 0.99 | 0.27, 3.59 | 0.98 |
| Chloride ≥ 110 mEq/L |  |  |  |  |  |  |
| Potassium ≥ 5.5 mEq/L | 2.39 | 0.96, 5.90 | 0.05 | 1.25 | 0.38, 4.12 | 0.70 |
| Sodium ≥ 148 mEq/L | 3.08 | 1.42, 6.68 | 0.00 | 1.53 | 0.53, 4.36 | 0.42 |
| Sodium <134 mEq/L | 0.68 | 0.26, 1.74 | 0.42 | 0.35 | 0.11, 1.08 | 0.06 |
| Fluid balance (mL) | 1.35 | 1.14, 1.61 | 0.00 | 1.15 | 0.87, 1.50 | 0.28 |
| Acute kidney injury | 3.21 | 1.93, 5.33 | 0.00 | 1.72 | 0.72, 4.11 | 0.22 |
| *Medications* | | | | | | |
| MRC-ICU mean (SD) | 1.06 | 1.04, 1.09 | 0.00 | 1.05 | 0.98, 1.13 | 0.14 |
| Vasopressor use in first 24 hours | 2.24 | 1.37, 3.67 | 0.00 | 0.60 | 0.24, 1.47 | 0.26 |
| Use of continuous infusions | 2.04 | 1.25, 3.33 | 0.00 | 1.07 | 0.50, 2.31 | 0.84 |
| Number of continuous infusions | 1.05 | 0.98, 1.13 | 0.14 | 0.87 | 0.73, 1.04 | 0.14 |
| *Data are presented as n (%) or mean ± standard deviation (SD) unless otherwise stated.*  *Albumin and lactate are not presented due to missingness exceeding 30%.* | | | | | | |

**Supplemental Table 2.** Stepwise regression for final model to predict presence of fluid overload

| **Variable** | **Odds Ratio** | **95% Confidence Interval** | **p-value** |
| --- | --- | --- | --- |
| Admission Diagnosis-Sepsis/septic shock | 3.52 | 1.64, 7.54 | 0.00 |
| Male | 0.44 | 0.25, 0.77 | 0.00 |
| SOFA at 24 hours | 1.31 | 1.20, 1.42 | 0.00 |
| Sodium ≥ 148 mEq/L | 2.11 | 0.87, 5.11 | 0.09 |
| Sodium <134 mEq/L | 0.41 | 0.14, 1.13 | 0.08 |
| Bicarbonate < 22 mEq/L | 0.51 | 0.29, 0.90 | 0.02 |
| Bicarbonate > 29 mEq/L | 0.29 | 0.11, 0.77 | 0.01 |
| SOFA-Sequential Organ Failure Assessment, OR- odds ratio, CI- confidence interval | | | |

**Supplemental Table 3.** Full regression model for amount of fluid overload at 72 hours

|  | **Univariate** | | | | **Multivariate** | | |
| --- | --- | --- | --- | --- | --- | --- | --- |
| **Variable** | **Odds Ratio** | **95% CI** | **p-value** | | **Odds Ratio** | **95% CI** | **p-value** |
| **ICU Baseline** | | | | | | | |
| Age ≥ 65 years old; mean (SD) | 0.00 | -0.01, 0.00 | | 0.08 | 0.00 | -0.01, 0.00 | 0.02 |
| Sex (male) | -0.01 | -0.01, 0.00 | | 0.00 | 0.00 | -0.01, 0.00 | 0.00 |
| *Relevant chronic conditions* | | | | | | | |
| Chronic kidney disease | -0.00 | -0.03, 0.01 | | 0.50 | -0.01 | -0.03, 0.00 | 0.22 |
| Heart failure | -0.00 | -0.0, 0.01 | | 0.62 | 0.01 | 0.00, 0.03 | 0.13 |
| Admission to medical ICU | 0.00 | 0.00, 0.01 | | 0.12 | 0.00 | 0.00, 0.01 | 0.77 |
| *Primary ICU Admission Diagnosis* | | | | | | | |
| Cardiac | -0.01 | -0.02, 0.00 | | 0.00 | -0.01 | -0.02, 0.00 | 0.02 |
| Hepatic | 0.00 | -0.02, 0.03 | | 0.83 | 0.00 | -0.03, 0.0 | 0.46 |
| Pulmonary | 0.00 | 0.00, 0.01 | | 0.59 | 0.00 | -0.01, 0.01 | 0.91 |
| Sepsis/septic shock | 0.03 | 0.01, 0.04 | | 0.00 | 0.01 | 0.00, 0.02 | 0.02 |
| Trauma | 0.00 | -0.01, 0.01 | | 0.82 | -0.01 | -0.03, 0.00 | 0.03 |
| **24 hours after ICU admission** | | | | | | | |
| *Severity of illness, using worst values recorded* | | | | | | | |
| APACHE II Score | 0.00 | 0.00, 0.00 | | 0.00 | 0.00 | 0.00, 0.00 | 0.03 |
| SOFA Score | 0.00 | 0.00, 0.00 | | 0.00 | 0.00 | 0.00, 0.00 | 0.01 |
| *Supportive care devices* | | | | | | | |
| Renal replacement therapy | -0.00 | -0.03, 0.02 | | 0.73 | -0.05 | -0.07, -0.02 | 0.00 |
| Invasive mechanical ventilation | 0.02 | 0.02, 0.03 | | 0.00 | -0.00 | -0.01, 0.00 | 0.17 |
| *Laboratory values (serum) and flowsheet values* | | | | | | | |
| Bicarbonate < 22 mEq/L | -0.02 | -0.03, -0.01 | | 0.00 | -0.00 | -0.01, 0.00 | 0.10 |
| Bicarbonate > 29 mEq/L | -0.03 | -0.04, -0.02 | | 0.00 | -0.01 | -0.02, 0.00 | 0.03 |
| Creatinine ≥ 1.5 mg/dL | 0.02 | 0.01, 0.03 | | 0.00 | 0.0019 | -0.01, 0.02 | 0.84 |
| Chloride ≥ 110 mEq/L | 0.02 | 0.01, 0.03 | | 0.00 | 0.00 | 0.00, 0.01 | 0.21 |
| Potassium ≥ 5.5 mEq/L | 0.02 | 0.00, 0.03 | | 0.01 | 0.00 | -0.01, 0.01 | 0.87 |
| Sodium ≥ 148 mEq/L | 0.03 | 0.02, 0.05 | | 0.00 | 0.0154 | 0.00, 0.031 | 0.05 |
| Sodium <134 mEq/L | 0.00 | -0.01, 0.00 | | 0.66 | -0.01 | -0.02, 0.00 | 0.05 |
| Fluid balance (mL) | 0.00 | 0.00, 0.00 | | 0.00 | 0.00 | 0.00, 0.00 | 0.03 |
| Acute kidney injury | 0.03 | 0.01, 0.04 | | 0.00 | 0.01 | 0.00, 0.02 | 0.05 |
| *Medications* | | | | | | | |
| MRC-ICU mean (SD) | 0.00 | 0.00, 0.00 | | 0.00 | 0.00 | 0.00, 0.00 | 0.01 |
| Vasopressor use in first 24 hours | 0.00 | 0.00, 0.00 | | 0.00 | 0.00 | 0.00, 0.00 | 0.01 |
| Use of continuous infusions | 0.02 | 0.01, 0.02 | | 0.00 | 0.00 | 0.00, 0.01 | 0.13 |
| Number of continuous infusions | 0.00 | 0.00, 0.00 | | 0.00 | 0.00 | 0.00, 0.00 | 0.05 |
| *Data are presented as n (%) or mean ± standard deviation (SD) unless otherwise stated.*  *Albumin and lactate are not presented due to missingness exceeding 30%.* | | | | | | | |

**Supplemental Table 4.** Stepwise regression for final model to predict fluid overload amount

| **Variable** | **Estimate** | **95% CI** | **p-value** |
| --- | --- | --- | --- |
| Fluid balance at 24 hr (mL) | 0.00 | 0.00, 0.00 | 0.01 |
| Sodium ≥ 148 mEq/L | 0.02 | 0.00, 0.03 | 0.00 |
| Sodium <134 mEq/L | -0.00 | -0.02, 0.00 | 0.10 |
| Sex (Male) | -0.00 | -0.01, 0.00 | 0.00 |
| Admission Diagnosis-Trauma | -0.01 | -0.03, 0.00 | 0.02 |
| Admission Diagnosis-Sepsis/septic shock | 0.01 | 0.00, 0.03 | 0.00 |
| Admission Diagnosis-Cardiac | -0.01 | -0.02, 0.00 | 0.00 |
| Age (>=65) | -0.00 | -0.01, 0.00 | 0.00 |
| SOFA at 24 hours | 0.00 | 0.00, 0.00 | 0.00 |
| Bicarbonate < 22 mEq/L | -0.01 | -0.02, 0.00 | 0.06 |
| Bicarbonate > 29 mEq/L | -0.01 | -0.02, 0.00 | 0.00 |
| APACHE II at 24 hours | 0.00 | 0.00, 0.002 | 0.00 |
| APACHE II- Acute Physiology and Chronic Health Evaluation, SOFA- sequential organ failure assessment | | | |

**Supplemental Table 5.** Performance of fluid overload prediction models for amount of fluid overload

| **Variable** | **Full Regression** | **Stepwise Selected Regression** | **MRC-ICU Regression** | **Random Forest** | **Support Vector Machine** | **XGBoost** |
| --- | --- | --- | --- | --- | --- | --- |
| Mean Squared Error | 0.002 | 0.002 | 0.002 | 0.001 | 0.002 | 0.002 |

**Supplemental Figure 1.** Consort diagram

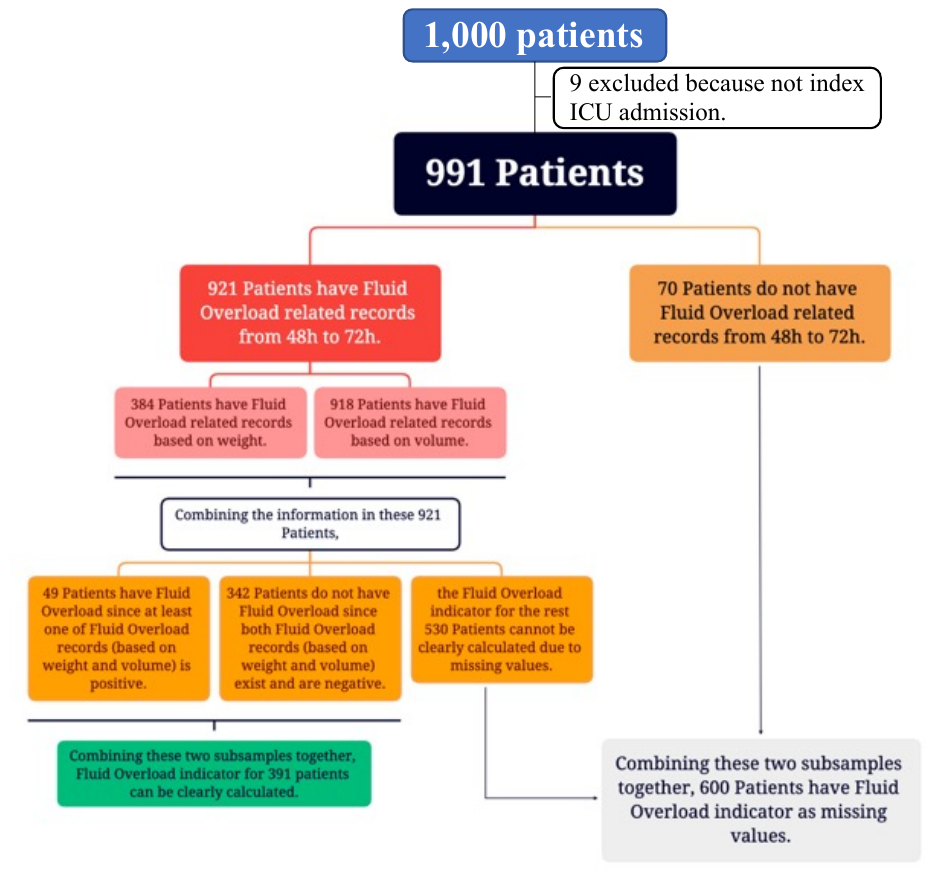

**Supplemental Figure 2.** AUROC Curve for Fluid Overload at 72 hours

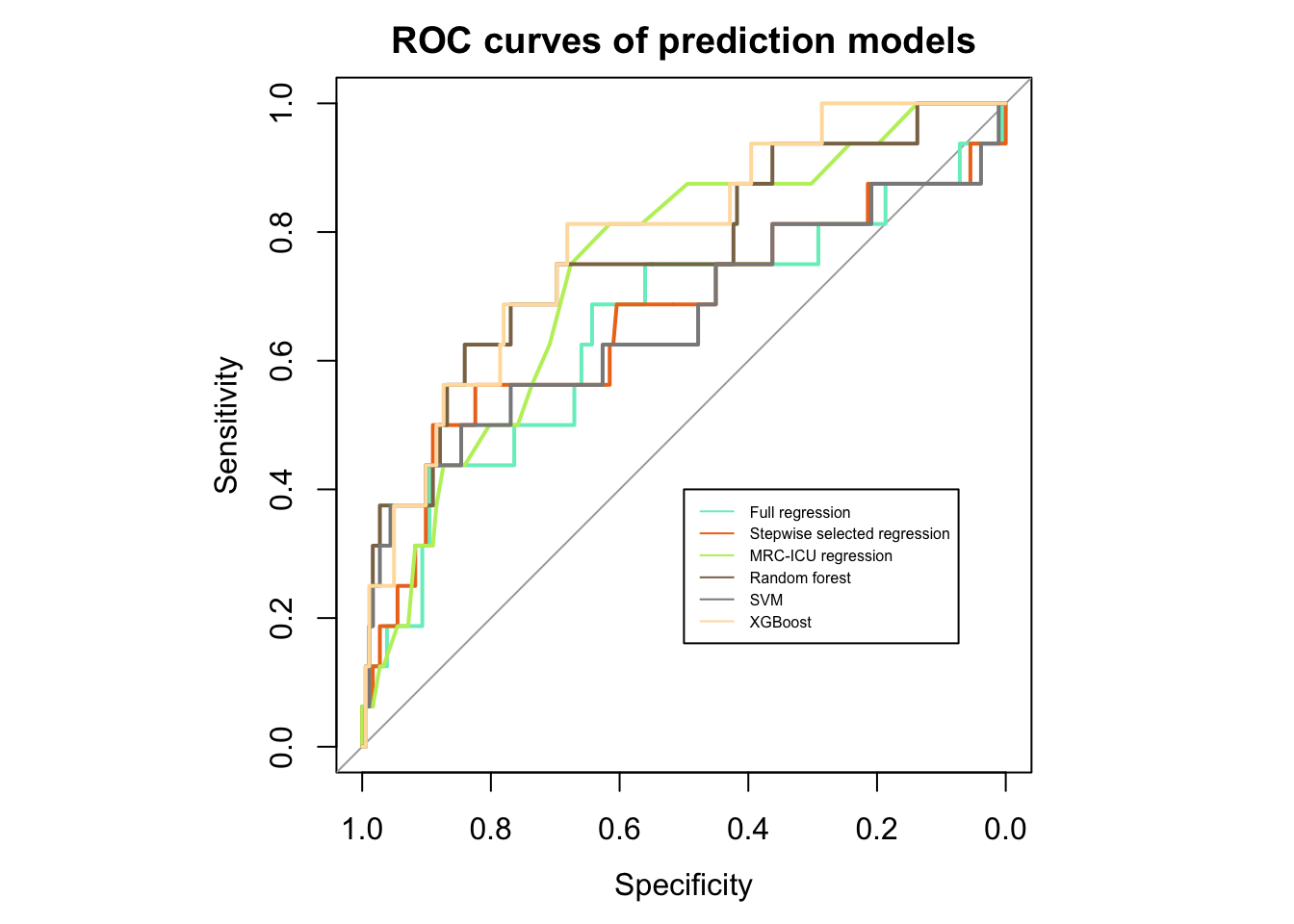

**Supplemental Figure 3.** Most common features for presence of fluid overload prediction with Random Forest imputations

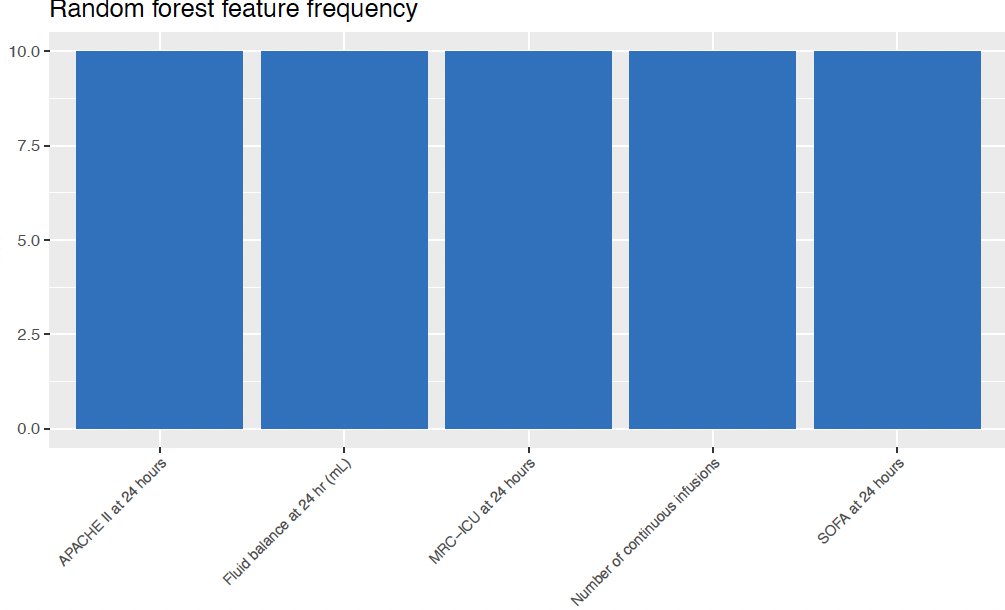

**Supplemental Figure 4.** Most common features for presence of fluid overload prediction with Support Vector Machine imputations

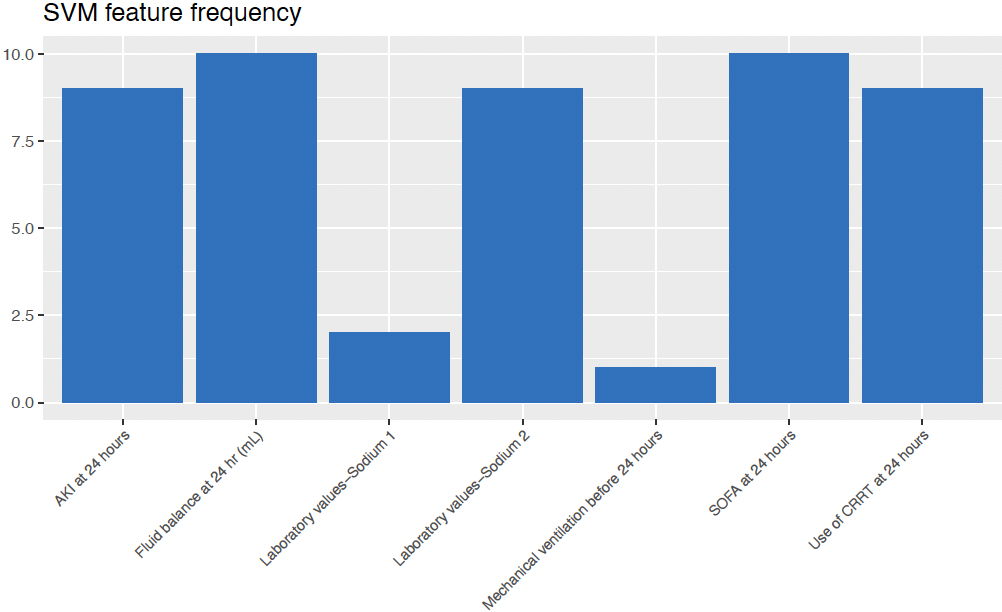

**Supplemental Figure 5.** Most common features for amount of fluid overload prediction with XGBoost imputations

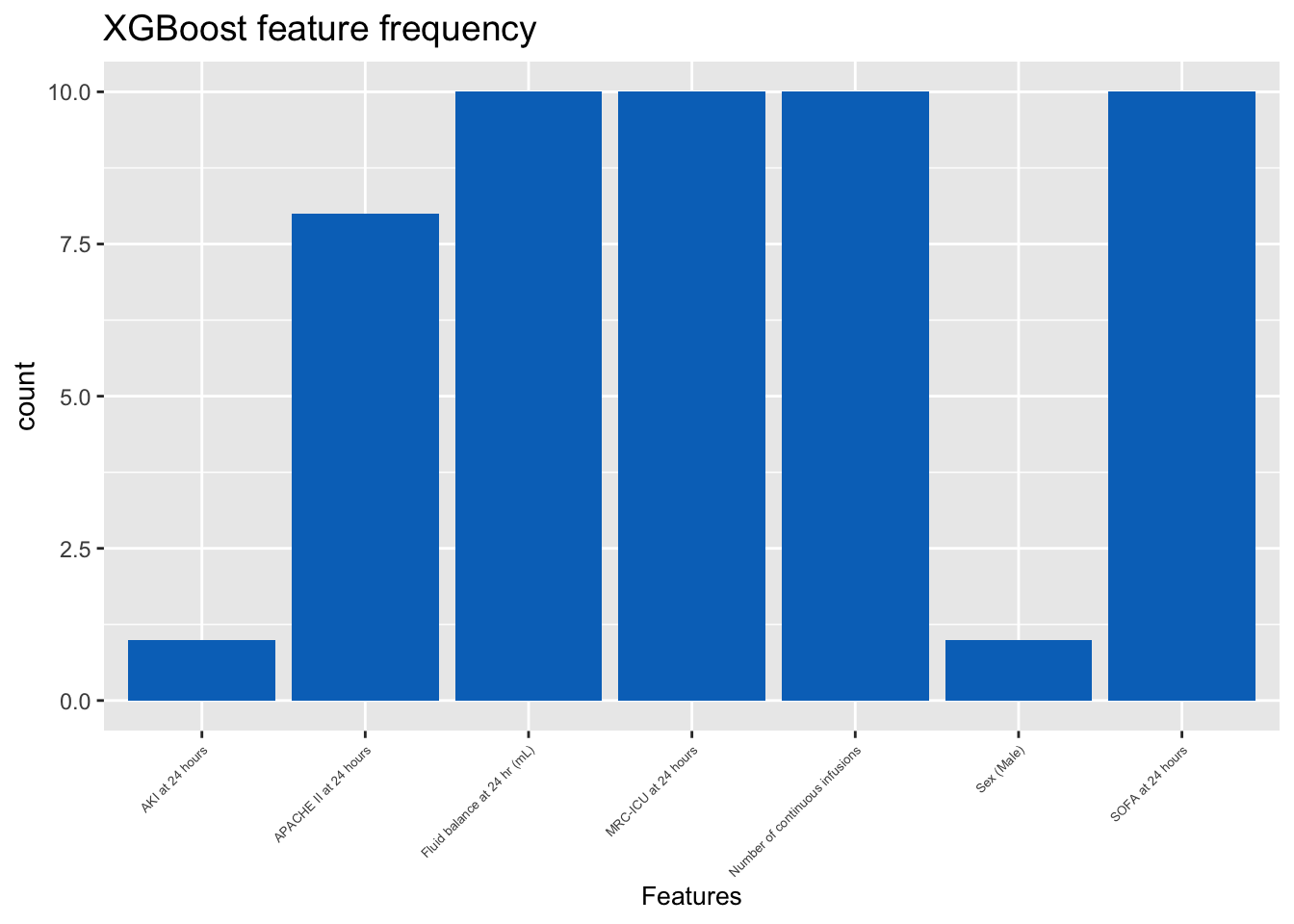

**Supplemental Figure 6.** Most common features for amount of fluid overload prediction with Random Forest imputations

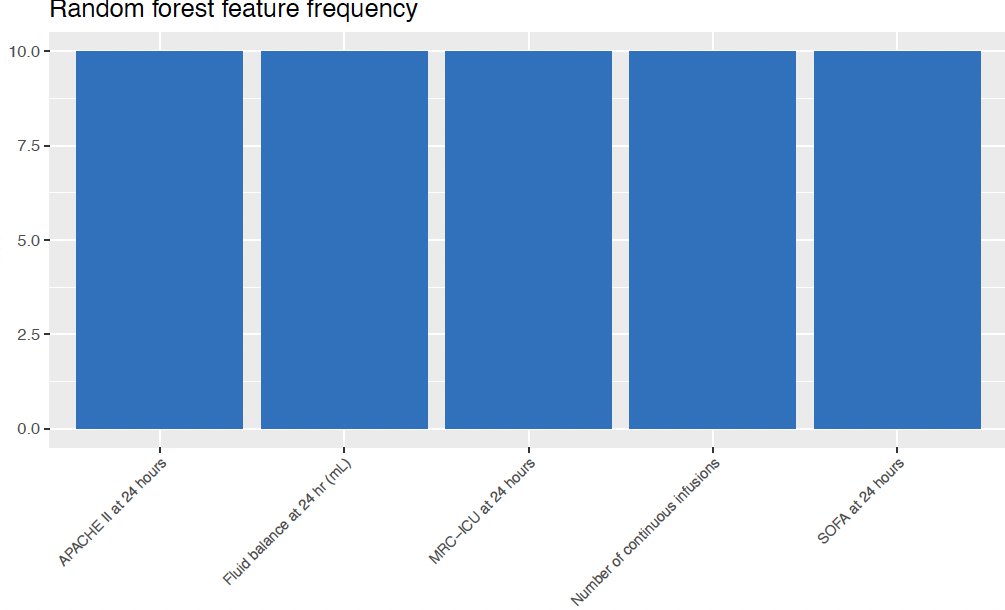

**Supplemental Figure 7.** Most common features for amount of fluid overload prediction with Support Vector Machine imputations

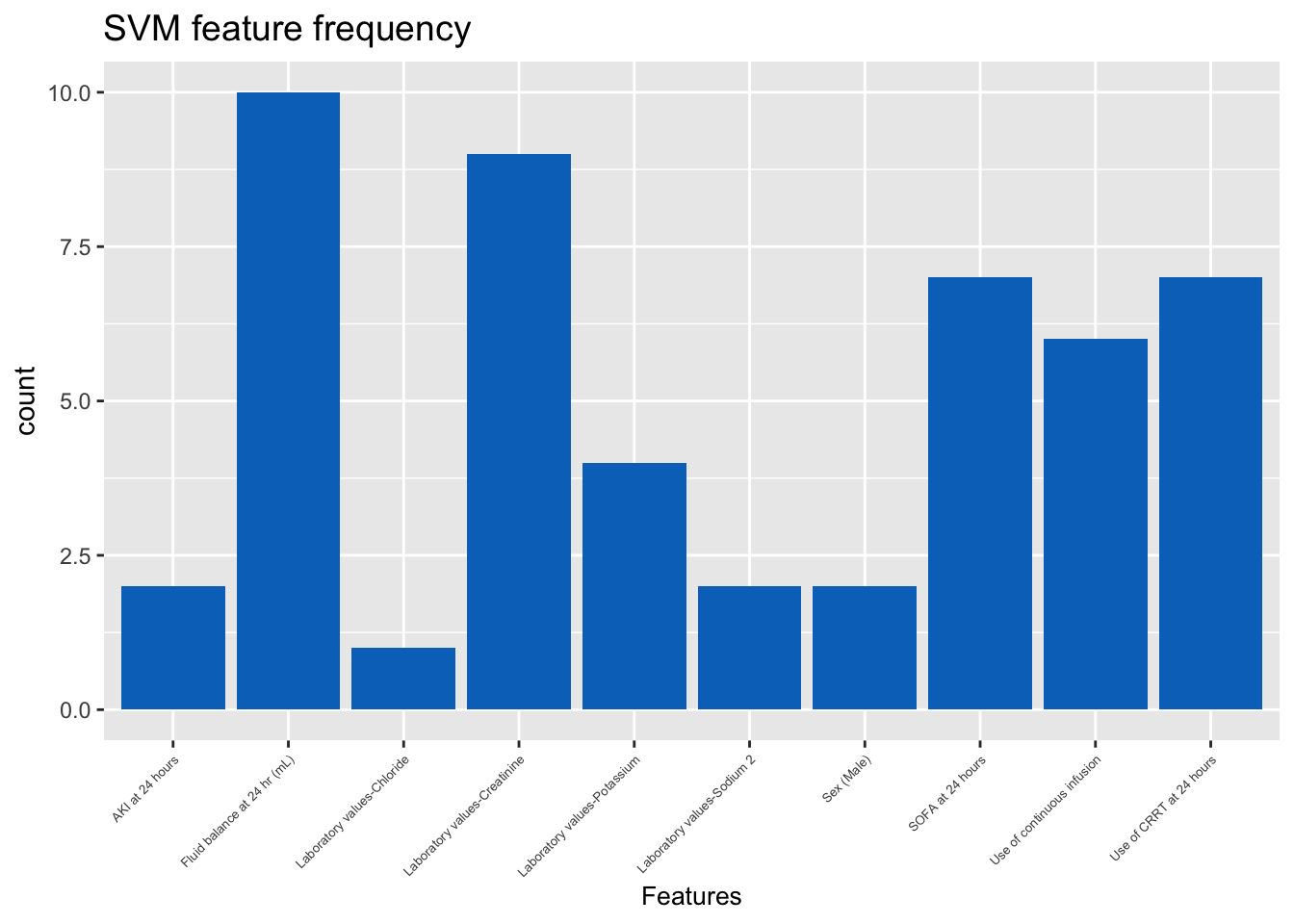
